# Trade-offs in emergency transport protocols for access to hip fracture management: a geospatial analysis of selective versus standard transfer in Ontario long-term care

**DOI:** 10.64898/2026.04.12.26350713

**Authors:** Nicholas J. Yee, Tianhui Chen, Yu Qing Huang, Cari M. Whyne, Mansur Halai

## Abstract

**Objectives:** For suspected hip fractures, prehospital protocols directing patients to an orthopaedic centre rather than the nearest emergency department (ED) could reduce time-to-surgery but may impact EMS travel burden. This study evaluates the impact of transfer protocols by quantifying transport to hospitals from long term care (LTC) facilities across Ontario.

**Methods:** A retrospective cross-sectional analysis of all Ontario LTC facilities and hospitals was performed. Two protocols were modeled: *standard transfer* to the nearest ED with subsequent transfer if required, and *selective transfer* based on Collingwood Hip Fracture Rule prehospital screening^1^directly to the nearest orthopaedic services (orthoED). Median one-way travel distances were calculated from Google Maps.

**Results:** In Ontario, 15.4% of LTC residents require hospital destination decisions because their nearest ED lacks orthopaedic services; for these facilities, median distances were 2.7km to the ED and 36.0km to the orthoED. Among the 52 LTC facilities where selective transfer was distance-optimal, it substantially reduced travel for patients with hip fracture (31.1km vs 49.6km; *P*<.01) while only modestly increasing travel for patients without hip fracture. Where standard transfer was distance-optimal, little travel difference was noted for patients with hip fracture, however false positive screened patients traveled significantly further to an orthoED. Greatest negative consequences of selective transfer lie in the 1.3% of residents living farthest (>100km) from an orthoED.

**Conclusions:** EMS direct transportation to hospitals with orthopaedics may improve hip fracture care but can increase EMS burden due to patients identified falsely as having a hip fracture, particularly in remote communities.

## Introduction

Hip fractures are common orthopaedic injuries requiring hospital admission and timely surgical management. The estimated lifetime risk is 7.3% for women and 6.2% for men^2^. These injuries carry substantial morbidity and mortality, with reported mortality rates of 23.5% following a first hip fracture and up to 66.5% after a second^3,4^. In 2022, hip fracture incidence was 574 per 100,000 among Canadian women and 363 per 100,000 among men; in Ontario, the rates were 534 and 351 per 100,000, respectively^5^. The mean one-year cost per case is $26,527 CAD, and the total national economic burden was $650 million in 2001, projected to rise to $2.4 billion by 2041, making it an important concern for healthcare systems^6^.

In 2021, 17.8% of Canadians lived in rural areas, which have a higher and faster-growing senior population compared to urban centers.^7,8^ Between 2009 and 2021, the proportion of those aged 65+ increased from 15% to 23.3% in rural areas, versus 13% to 18.2% in urban areas.^7,9^ This demographic shift raises concerns as most orthopaedic surgical resources are concentrated in urban hospitals, potentially worsening future healthcare access and patient outcome disparities.

The standard of care for hip fractures is operative management within 48 hours, as delays in time-to-surgery are associated with increased mortality, postoperative complications, and prolonged hospital stay^10–14^. Rural patients typically travel to their nearest emergency departments for initial assessments, which may not offer orthopaedic surgery services, and thus requiring subsequent inter-hospital transfer when surgical management is needed. In Ontario, transferred patients had a time-to-surgery of 93 hours compared to 44 hours for non-transferred patients; with the delays mainly occurring prior to the inter-hospital transfer^15^. Transfer status itself has been independently associated with worse outcomes and increased healthcare utilization^10,15–18^.

Emergency medical services (EMS) protocols typically prioritize transport to the nearest emergency department (ED) for initial evaluation. While this standard transfer approach ensures rapid access to emergency care, it may result in secondary inter-hospital transfers for patients who require operative management. Direct transport of all suspected hip fracture patients to hospitals with orthopaedic services is ill-advised as only 25.7% of suspected hip fracture patients are ultimately diagnosed with a fracture^1^. Recent literature has explored the use of prehospital screening tools, such as the Collingwood Hip Fracture Rule (CHFR), to improve screening of patients with suspected hip fracture and enable direct transport to hospitals with orthopaedic services^1^. These strategies have the potential to reduce interhospital transfers and time-to-surgery but raise concerns that direct transport to orthopaedic centers may increase travel distances and EMS resource utilization as many patients do not have hip fractures and could be mistakenly screened positive.

Given these considerations, there is a need to quantify how different EMS transport protocols influence access to orthopaedic care and travel burden, especially for EMS servicing long-term care (LTC) facilities. LTC facilities are residential homes that provide 24-hour supervised health, personal, and social support to individuals whose needs cannot be met independently. LTC facilities have a high density of residents at high risk for hip fractures and are located across the province. Understanding these trade-offs are essential for informing regionalized trauma protocols that balance timely surgical care with efficient use of EMS resources.

The objective of this study is to perform a province-wide geospatial analysis of Ontario LTC facilities and hospital locations to compare travel distances under standard transfer (transport to the nearest ED) and selective transfer (direct transport to an hospital with ED and orthopaedic services based on prehospital screening). We aim to identify LTC facilities where paramedic triage decisions are required, to determine geographic conditions that selective transfer reduce expected travel burden, and to characterize regions in Ontario where selective transfer may increase EMS transport burden.

## Methods

### Study Design

This study was a retrospective cross-sectional geospatial analysis of Ontario LTC facilities and hospital locations. We compared emergency medical services (EMS) travel distances under two triage protocols for patients with suspected hip fracture: a standard transfer protocol and a selective transfer protocol.

### Setting and Participants

All publicly listed LTC facilities and general hospitals in Ontario were identified using publicly available data from the Government of Ontario websites (LTCs: https://www.ontario.ca/page/long-term-care-ontario; hospitals: https://www.ontario.ca/page/general-hospital-locations). Data were collected in October 2023. Hospitals were classified according to the presence of an emergency department and orthopaedic services based on information obtained from official hospital websites. Only hospitals with emergency departments (ED) or both emergency departments and orthopaedic services (orthoED) were included. All LTC facilities had waitlists. As such, the number of beds was used as a proxy for the number of residents at each LTC facility.

The primary analysis focused on LTC facilities where the nearest ED did not also provide orthopaedic services (i.e., the nearest ED was not an orthoED), as these represent facilities where EMS paramedics must determine the most appropriate destination hospital. This study used publicly available institutional-level data.

### Prehospital triaging protocol

Two EMS transport strategies were modeled. Under the standard transfer protocol, patients were transported from the LTC facility to the nearest ED, with subsequent inter-hospital transfer to the hospital’s nearest orthoED for hip fracture patients. Under the selective transfer protocol, patients with a positive prehospital screening test for hip fractures were transported directly to the nearest orthoED, whereas those with a negative screen were transported to the nearest ED. False negatives were modelled to have an inter-hospital transfer to the nearest orthoED.

The screening test was modeled using a 85% sensitivity and 95% specificity with a 25% hip fracture incidence, consistent with a previously published CHFR prehospital screening tool for hip fractures^1^. Travel distances between LTC facilities and hospitals were calculated using the Google Maps Distance Matrix API (Google LLC, California, USA). For each LTC facility, the shortest one-way distance to the nearest ED, the distance to the nearest orthoED, and the indirect distance from the LTC to the ED and subsequently to the orthoED were calculated.

The expected patients’ travel distances under each triage protocol were derived as weighted averages based on the probability of true positive, false positive, true negative, and false negative screening outcomes, incorporating both the hip fracture incidence and the diagnostic performance of the CHFR prehospital screening tool. Map visualizations used GeoPandas (v1.1.3), contextily (v1.7.0), and OpenStreetMap with Mapnik (OpenStreetMap contributors, licensed under the Open Data Commons Open Database License).

### Outcome Measures

The primary outcome was the one-way EMS travel distance to ED/orthoED for LTC facilities where the destination hospital is a required triaging decision. The expected travel distance for patients with hip fracture, without hip fracture, and for the weighted average patient was reported as a median and interquartile range (IQR)^1^. Secondary outcomes included the identification of Ontario LTC facilities that would have increased travel distances by implementing a direct transport to orthoED triaging protocol. Travel distance was used instead of travel time as it is less traffic-dependent and more reproducible.

### Statistical Analysis

Comparisons between expected travel distances under the standard and selective transfer strategies were performed using the Wilcoxon signed-rank test. Statistical analyses were conducted using Python (v3.14.3) with Pandas (v3.0.1), NumPy (v2.4.3) and Pingouin (v0.6.1). A two-sided significance threshold of α = 0.01 was used.

## Results

A total of 626 LTC facilities comprising 79,710 licensed beds for LTC residents and 171 hospitals with emergency departments were identified across Ontario, including 96 hospitals also with orthopaedic services. All LTC facilities were accessible by road to at least one emergency department. Of these, 148 LTC facilities (representing 15.4% of LTC residents) had a nearest ED that did not provide orthopaedic services and were included in the comparative analysis of triage protocols (Figure 1). The remaining LTC facilities, where the nearest ED also provided orthopaedic services, were excluded from subsequent analyses as both transfer strategies would result in the same destination.

**Figure 1:**
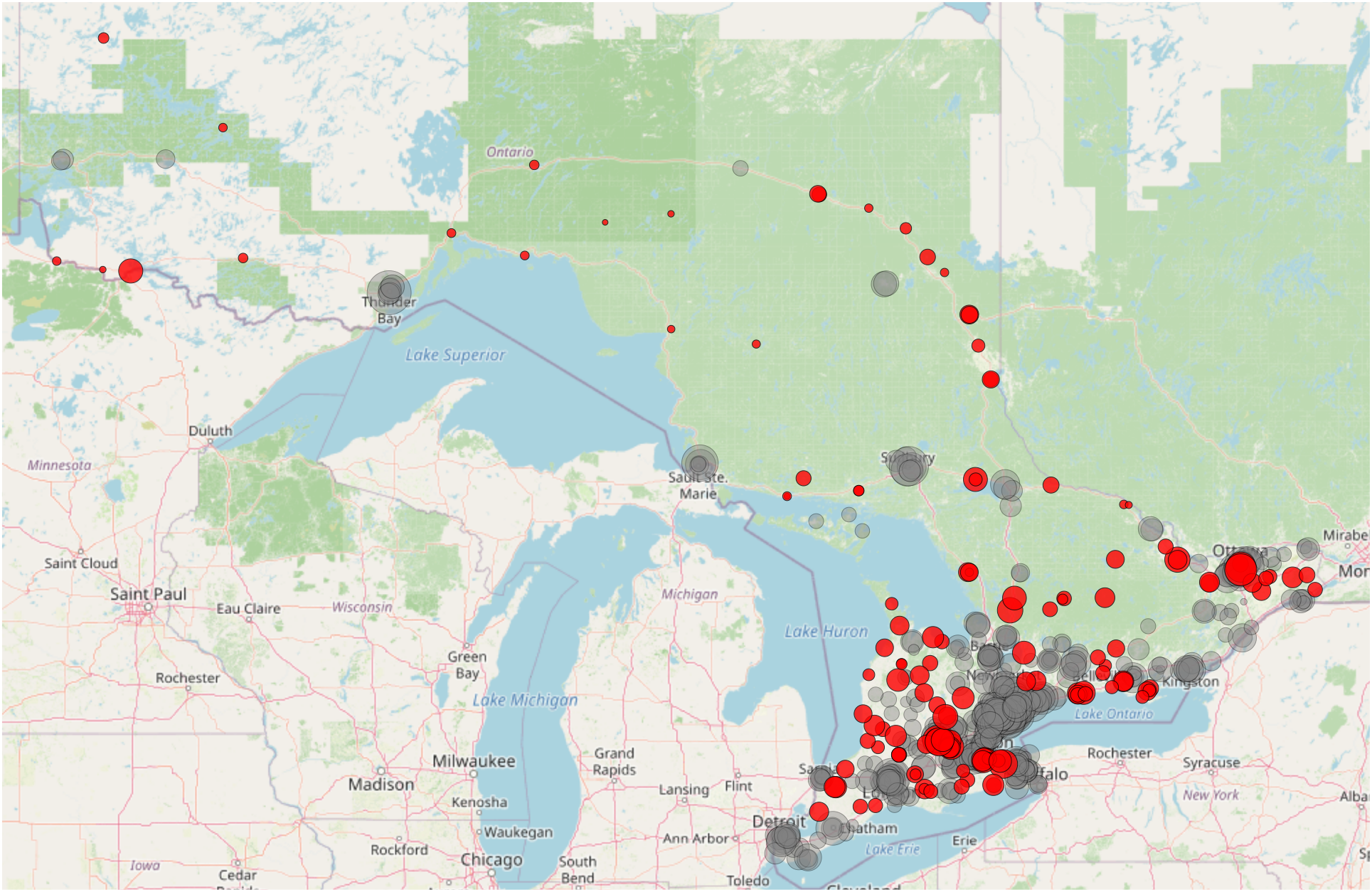
Distribution of Ontario LTC facilities. Red points indicate facilities where a decision on the destination hospital for the hip trauma patient is required (i.e., the nearest hospital with an emergency department does not provide orthopaedic services). Gray points indicate facilities where the nearest hospital with an emergency department also offers orthopaedic services. Point size is proportional to the number of beds at the LTC facility. All publicly listed Ontario LTC facilities are shown.

Among LTC facilities requiring a triage decision, the median distance to the nearest ED was 2.7 [0.7 – 8.6] km, whereas the median distance to the nearest orthoED was 36.0 [23.0 – 68.0] km. The median indirect travel distance from LTC to ED followed by transfer to their nearest orthoED was 44.4 [26.2 – 76.3] km. Substantial variability was observed in the relative distances to ED and orthoED across LTC facilities (Figure 2).

**Figure 2:**
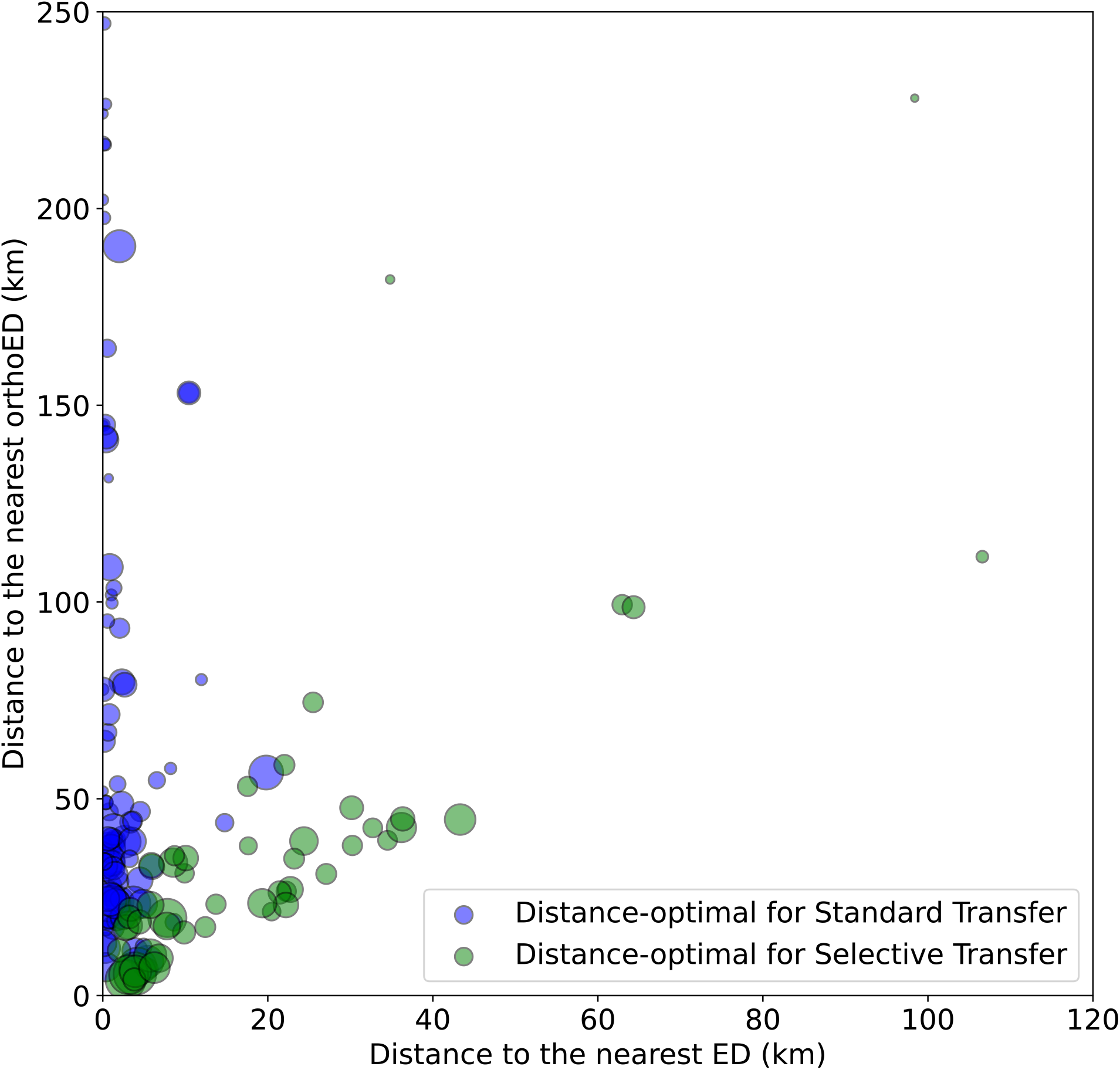
Scatter plot of long-term care (LTC) facilities showing the distance to their nearest emergency department (ED) and their nearest hospital with an emergency department and orthopaedic service (orthoED). Point size is proportional to the number of beds at the LTC facility. LTC facilities where their nearest ED is their nearest orthoED were excluded.

Under universally applied standard transfer and selective transfer protocols, the travel distance for the average patient was 14.0 [8.6 – 30.6] km and 14.9 [8.5 – 29.7] km, respectively. For hip fracture patients, they would travel 44.4 [26.2 – 76.3] km and 37.9 [23.7 – 68.1] km, respectively. Non-hip fracture patients would travel 2.7 [0.7 – 8.6] km and 5.8 [3.1 – 11.3] km, respectively.

96 LTC facilities (representing 9.1% of the Ontario LTC residents) would minimize their average patient travel distance through the standard transfer protocol (standard: 12.4 [8.3 – 25.9] km; selective: 13.5 [8.5 – 28.7] km), whereas 52 facilities (6.3% of the residents) were better served by the selective transfer protocol (standard: 21.8 [9.1 – 36.0] km; selective: 19.8 [8.7 – 33.0] km). Facilities favoring standard transfer were generally characterized by shorter distances to ED and further distances to orthoED (Figure 2, Table 1).

**Table 1:** Transport distances from long-term care (LTC) facilities to hospital destinations, stratified by facilities where standard versus selective transfer is distance-optimal for the average patient. Average patient travel distance is a weighted average using a 25% hip fracture incidence and screening test sensitivity of 85% and specificity of 95%. Distances are shown for transport to the nearest emergency department (ED), directly to the nearest hospital with an emergency department and orthopaedic service (orthoED), and via ED followed by interhospital transfer to orthoED. Values are reported as a median and interquartile range.

|  | LTC Favoring Standard Transfer (km) | LTC Favoring Selective Transfer (km) |
| --- | --- | --- |
| LTC to the nearest ED | 1.1 (0.4 - 2.5) | 15.6 (5.8 - 25.9) |
| LTC to the nearest orthoED | 39.9 (24.8 - 91.6) | 26.7 (17.6 - 40.2) |
| LTC to the nearest ED then to the nearest orthoED | 40.7 (26.4 - 92.8) | 49.6 (25.2 - 64.7) |

Among LTC facilities where standard transfer was distance-optimal for the average patient, implementing a selective transfer resulted in slightly shorter travel distances for patients with hip fracture (40.0 [24.9–91.8] km vs 40.7 [26.4–92.8] km; *P*<.01), but substantially increased travel distances for patients without hip fracture due transferring false positive screened patients to the further orthoED (4.3 [2.6–7.3] km vs 1.1 [0.4–2.5] km; *P*<.01). This led to a higher overall average travel distance with selective transfer for those LTC facilities (13.5 [8.5–28.7] km vs 12.4 [8.3–25.9] km; *P*<.01; Table 2). Conversely, among LTC facilities where selective transfer was distance-optimal, selective transfer substantially reduced travel distances for patients with hip fracture (31.1 [19.2–43.5] km vs 49.6 [25.2–64.7] km; *P*<.01) while only modestly increasing travel distances for patients without hip fracture (16.4 [6.2–27.4] km vs 15.6 [5.8–25.9] km; *P*<.01), resulting in a lower average travel distance (19.8 [8.7–33.0] km vs 21.8 [9.1–36.0] km; *P*<.01).

**Table 2:** Expected emergency medical transport distances under standard versus selective transfer protocol, stratified by longterm care (LTC) facilities where each strategy is distance-optimal for the average patient. Average patient travel distance is a weighted average using a 25% hip fracture incidence and screening test sensitivity of 85% and specificity of 95%. Distances are reported for patients with hip fracture, without hip fracture, and the weighted average patient. Values are presented as a median and interquartile range.

| Protocol | LTC favoring standard transfer (km) |  |  | LTC favoring selective transfer (km) |  |  |
| --- | --- | --- | --- | --- | --- | --- |
|  | Hip fracture | No hip fracture | Average patient | Hip fracture | No hip fracture | Average patient |
| Standard | 40.7 (26.4 - 92.8) | 1.1 (0.4 - 2.5) | 12.4 (8.3 - 25.9) | 49.6 (25.2 - 64.7) | 15.6 (5.8 - 25.9) | 21.8 (9.1 - 36.0) |
| Selective | 40.0 (24.9 - 91.8) | 4.3 (2.6 - 7.3) | 13.5 (8.5 - 28.7) | 31.1 (19.2 - 43.5) | 16.4 (6.2 - 27.4) | 19.8 (8.7 - 33.0) |

Among LTC facilities located more than 100 km from the nearest orthoED, 24 facilities (1.3% of residents) were identified, most of which were located north of Lake Huron (Figure 3). Their travel distance to ED was 0.5 [0.2–3.1] km whereas travel to the orthoED was 156 [141.7–205.7] km. As a result, the standard transfer protocol was more frequently distance-optimal for the average patient (48.8 [36.2–54.8] km) vs selective strategy (51.9 [41.6 – 62.9] km) despite the selective strategy’s potential reduction in interhospital transfers. Patients sent to orthoED would endure significantly longer transportation which raises important considerations of patient preference in their trauma management.

**Figure 3:**
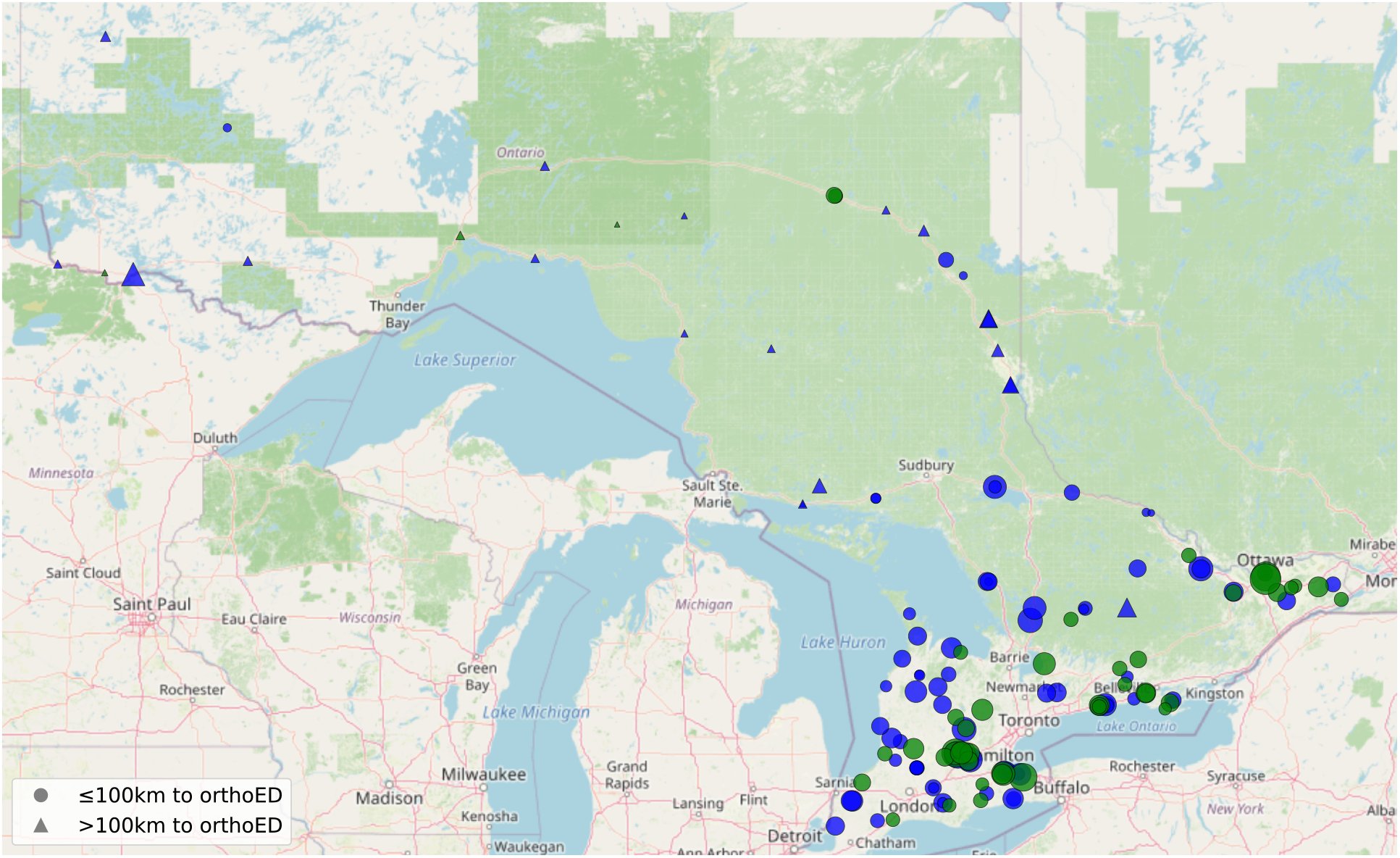
Distribution of Ontario long-term care (LTC) facilities. LTC facilities that need a triaging decision on the destination hospital are shown. LTC facilities located >100km from their nearest hospital with an emergency department and orthopaedic service (orthoED) are shown with a triangle. Blue points indicate facilities where the optimal triaging protocol based on minimizing travel distance is to always present to ED first and perform an interhospital transfer for hip fracture patients (standard transfer); green points indicate facilities that would benefit from following the selective transfer protocol using a prehospital screening tool to send positive screened patients directly to orthoED. Point size is proportional to the number of beds at the LTC facility.

## Discussion

This study presents a province-wide geospatial analysis of the implementation of EMS transport protocols for suspected hip fracture among Ontario LTC residents. A substantial proportion of Ontario LTC facilities require triage decisions on their destination hospital because the nearest ED does not provide orthopaedic services. For these LTC residents with hip fractures, they may require an inter-hospital transfer, which is associated with increased rates of post-operative complications, including mortality^10,15–18^.

Diagnosis of hip fractures depend on comprehensive histories and physical exams and confirmation with diagnostic imaging. Older adult patients who fall and present with anterior groin pain, inability to weight bear, and shortened, externally rotated, and abducted limbs should be suspected to have a hip fracture. Gillette et al. proposes CHFR as the first clinical assessment screening tool aimed towards use by healthcare professionals in prehospital environments to triage suspected hip fracture patients for direct transport to a hospital with orthopaedic surgery services^1^.

Implementing a selective transfer protocol using a prehospital hip fracture screening tool would reduce inter-hospital transfers through selectively transferring positively screened patients directly to orthoED and bypassing their local ED. However, this could lead to greater EMS transportation burden due to an increase in averaged transport distance per patient, which may translate to higher local healthcare resource utilization. Given the low incidence of hip fractures among suspected hip trauma patients and despite the high specificity on prehospital screening tests, EMS would minimize medical transportation for most LTC facilities near an ED by using the standard protocol. This is disproportionately true for rural Ontario LTC as many facilities are located relatively near an ED but far from orthoED. Reducing inter-hospital transfer for these facilities by implementing selective transfer may further strain healthcare resources in rural communities. As such, identification of which LTC facilities may benefit from the selective protocol is critical prior to consideration of adoption.

Systematically reducing inter-hospital transfers by directly transporting patients to the appropriate hospital may benefit patient outcomes. Effective patient redirection relies on high specificity to minimize lengthy transfers for patients without hip fractures. The CHFR achieves an outstanding 95% specificity but our results shows that several Ontario EMS regions would increase their average transportation distance because of the low hip fracture incidence (25%) among older adult hip trauma patients requiring assessment^1^. The difference between these protocols would be much larger for a screening test specificity less than 95%. EMS have limited resources and medical directives for a selective transfer protocol like CHFR potentially strain regional EMS resources, despite its opportunity to improve patient outcomes.

Given the accuracy of diagnostic imaging, portable x-ray on the ambulance or in LTC facilities may allow early definitive diagnosis that would help with prehospital triaging. Point-of-care ultrasound (POCUS) is emerging as a viable alternative as it is affordable and does not use ionizing radiation. POCUS has a sensitivity of 95% (95% CI: 94 – 100%) and 70% (95% CI: 61 –79%) specificity^19–21^. However, its performance is highly dependent on access to trained sonographers and standard of care still requires x-ray, CT, or MRI for definitive diagnosis. POCUS may still improve triaging accuracy. The inclusion of point of care diagnostic imaging would additionally improve hip trauma assessment of non-verbal patients or patient unable to accurately communicate their symptoms (e.g., delirium, dementia, or confusion). However, implementing these technologies would require additional investment, training, and integration into existing EMS workflows.

Alternatively, optimizing inter-hospital transfer pathways, including expedited transfer protocols and coordination between referring and receiving centers, may reduce delays for patients initially transported to EDs without orthopaedic services. Several LTC facilities are located less than 1km away from ED. Fast-tracking patients to the radiology department in their ED also could reduce time-to-surgery^22,23^. Earlier diagnosis with prehospital or fast-tracked diagnostic imaging can reduce inter-hospital transfers and time before inter-hospital transfers.

Other strategies to improve the feasibility of prehospital screening may come from aligning regional healthcare restrictions, such as medical transport utilization, with province-wide healthcare metrics, such as time-to-surgery for hip fractures. Patients with hip fractures take up the largest number of beds in the ED^22^and delays in time-to-surgery for patients requiring interhospital transfers occur primarily before the transfer as patients wait for bed availability^15^. Introducing diagnostic imaging in prehospital environments or implementing selective transfer could initiate earlier discussions on bed availability with hospitals with orthopaedic services. In Canada, the basic hospital cost per day for surgical patients is $1,500 and patients requiring inter-hospital transfer tend to stay 7 days longer than directly transferred patients^15,24^. Funding structures to share cost savings with regional EMS may help with medical transportation budget concerns with implementing a selective transfer protocol or prehospital diagnostic imaging.

The study results suggest that a province-wide selective transfer protocol may not be optimal across all regions from an EMS resource utilization perspective. While selective transfer offers clear benefits to EMS resource utilization for a subset of LTC facilities, particularly those located closer to orthopaedic centers, standard transfer remains more efficient for many LTC facilities when considering the average patient. A region-specific approach to EMS triage may be warranted, incorporating geographic factors such as relative distances to ED and orthoED, as well as local EMS capacity. Considerations for expanding orthopaedic services in underserved regions could also reduce reliance on inter-hospital transfer and improve time-to-surgery. Additionally, system-level benefits beyond transport distance, including reduced duplication of investigations and improved coordination of care, should be considered when evaluating triage protocols.

This study represents a population-level analysis of all LTC facilities and hospitals in Ontario, incorporating real-world geographic data to model EMS transport patterns. The analysis accounts for both hip fracture incidence and screening test performance from the same region as the CHFR study, allowing for a realistic comparison of triage protocols. Additionally, the study identifies specific LTC facilities and regions where each transport protocol may be most appropriate, providing actionable insights for healthcare planning. There are also important limitations. This study is based on modeled transport distances and does not include patient-level outcomes such as their time-to-surgery, complications, or mortality. Travel distances were estimated using Google Maps and do not account for factors such as changes in traffic, weather, EMS response times and capacity, demographic changes, or seasonal volumes. The assumed hip fracture incidence and screening test characteristics may vary across populations and clinical settings. Hospital-level factors such as bed availability, operating room or post-operative ward capacity, and local transfer protocols were not included and may influence actual access to care. Additionally, hospital services were based on their websites which may lack detail such as whether orthopaedic services are offered daily or only certain days or weeks.

Future research should focus on prospective evaluation of selective transfer protocols, incorporating patient-level outcomes and cost analyses to better quantify their clinical and economic impact. Studies evaluating the implementation of prehospital screening tools in real-world EMS settings may want to include older adults living in the community in addition to LTC residents. The inclusion of patient perspectives on medical transport would add a valuable dimension for analysis. As the Canadian population is getting older, further work on exploring cost-effectiveness of hip fracture access to care would benefit our healthcare system, especially for our rural communities.

## Conclusions

In this province-wide geospatial analysis of Ontario LTC facilities, a substantial proportion of residents who fall with hip trauma were identified as requiring EMS triage decision on the destination hospital with ED or with ED and orthopaedic services. Selective transfer protocols would reduce inter-hospital transfers and shorten travel distances for patients with hip fracture, with potential to improve time-to-surgery and clinical outcomes. However, many LTC facilities would have increased EMS transport distances for the majority of patients without hip fracture. EMS in rural regions, particularly serving LTC facilities north of Lake Huron, would be disproportionately affected by implementing selective transfer as their orthoEDs are geographically distant with relatively nearby EDs.

This study highlights an important trade-off between patient-centered benefits and system-level resource utilization. A province-wide selective transfer strategy may not be optimal; instead, region-specific triage protocols that consider geographic access, EMS capacity, patient preference, and diagnostic accuracy may better balance efficiency and outcomes. Improvements in prehospital diagnostic capability, optimization of inter-hospital transfer pathways, and expansion of orthopaedic services in underserved areas may be avenues to improve access to timely hip fracture care.

## Data Availability

All data produced in the present study are available upon reasonable request to the authors.

## References

1. Gillette DM, Cheng O, Wilson A, Mantero R, Chisholm D, Feldman M. Screening tool for identification of hip fractures in the prehospital setting. OTA Int. 2021;4(4):e157. doi:10.1097/OI9.0000000000000157

2. Hopkins RB, Pullenayegum E, Goeree R, et al. Estimation of the lifetime risk of hip fracture for women and men in Canada. Osteoporos Int. 2012;23(3):921–927. doi:10.1007/s00198-011-1652-8

3. Berry SD, Samelson EJ, Hannan MT, et al. Second hip fracture in older men and women: the Framingham Study. Arch Intern Med. 2007;167(18):1971–1976. doi:10.1001/archinte.167.18.1971

4. Ioannidis G, Papaioannou A, Hopman WM, et al. Relation between fractures and mortality: results from the Canadian Multicentre Osteoporosis Study. CMAJ. 2009;181(5):265–271. doi:10.1503/cmaj.081720

5. Canadian Institute for Health Information. Hospitalized Hip Fracture Event. Accessed April 13, 2025. https://www.cihi.ca/en/indicators/hospitalized-hip-fracture-event?pageId=1114187

6. Wiktorowicz ME, Goeree R, Papaioannou A, Adachi JD, Papadimitropoulos E. Economic implications of hip fracture: health service use, institutional care and cost in Canada. Osteoporos Int. 2001;12(4):271–278. doi:10.1007/s001980170116

7. Dandy K, Bollman R. Seniors in rural Canada. Rural and Small Town Canada Analysis Bulletin. 2008;7:1–56.

8. Government of Canada SC. Population growth in Canada’s rural areas, 2016 to 2021. February 9, 2022. Accessed April 13, 2025. https://www12.statcan.gc.ca/census-recensement/2021/as-sa/98-200-x/2021002/98-200-x2021002-eng.cfm

9. Government of Canada SC. In the midst of high job vacancies and historically low unemployment, Canada faces record retirements from an aging labour force: number of seniors aged 65 and older grows six times faster than children 0-14. April 27, 2022. Accessed April 13, 2025. https://www150.statcan.gc.ca/n1/daily-quotidien/220427/dq220427a-eng.htm

10. Lawless AM, Narula S, D’Alessandro P, Jones CW, Seymour H, Yates PJ. Time to surgery and transfer-associated mortality for hip fractures in Western Australia. ANZ J Surg. 2020;90(9):1750–1753. doi:10.1111/ans.16115

11. Simunovic N, Devereaux PJ, Sprague S, et al. Effect of early surgery after hip fracture on mortality and complications: systematic review and meta-analysis. CMAJ. 2010;182(15):1609–1616. doi:10.1503/cmaj.092220

12. Moja L, Piatti A, Pecoraro V, et al. Timing matters in hip fracture surgery: patients operated within 48 hours have better outcomes. A meta-analysis and meta-regression of over 190,000 patients. PLoS One. 2012;7(10):e46175. doi:10.1371/journal.pone.0046175

13. Shiga T, Wajima Z, Ohe Y. Is operative delay associated with increased mortality of hip fracture patients? Systematic review, meta-analysis, and meta-regression. Can J Anaesth. 2008;55(3):146–154. doi:10.1007/BF03016088

14. Novack V, Jotkowitz A, Etzion O, Porath A. Does delay in surgery after hip fracture lead to worse outcomes? A multicenter survey. Int J Qual Health Care. 2007;19(3):170–176. doi:10.1093/intqhc/mzm003

15. Desai SJ, Patel J, Abdo H, Lawendy AR, Sanders D. A comparison of surgical delays in directly admitted versus transferred patients with hip fractures: opportunities for improvement? Can J Surg. 2014;57(1):40–43. doi:10.1503/cjs.002613

16. Hughes AJ, Brent L, Biesma R, Kenny PJ, Hurson CJ. The effect of indirect admission via hospital transfer on hip fracture patients in Ireland. Ir J Med Sci. 2019;188(2):517–524. doi:10.1007/s11845-018-1854-6

17. Lennox L, Myint PK, Baliga S, Farrow L. The Impact of Hospital Transfers on Surgical Delay and Associated Postoperative Outcomes for Hip Fracture Patients in Scotland: A Cohort Study. J Clin Med. 2024;13(9):2546. doi:10.3390/jcm13092546

18. Malik AT, Quatman CE, Phieffer LS, Ly TV, Jain N, Khan SN. Transfer status in geriatric hip fracture surgery-An independent risk factor associated with 30-day mortality, re-operations and complications. J Clin Orthop Trauma. 2019;10(Suppl 1):S65–S70. doi:10.1016/j.jcot.2019.01.025

19. Safran O, Goldman V, Applbaum Y, et al. Posttraumatic painful hip: sonography as a screening test for occult hip fractures. J Ultrasound Med. 2009;28(11):1447–1452. doi:10.7863/jum.2009.28.11.1447

20. Akimoto T, Kobayashi T, Maita H, Osawa H, Kato H. Initial assessment of femoral proximal fracture and acute hip arthritis using pocket-sized ultrasound: a prospective observational study in a primary care setting in Japan. BMC Musculoskelet Disord. 2020;21(1):291. doi:10.1186/s12891-020-03326-x

21. Cohen A, Li T, Greco J, et al. Hip effusions or iliopsoas hematomas on ultrasound in identifying hip fractures in the emergency department. The American Journal of Emergency Medicine. 2023;64:129–136. doi:10.1016/j.ajem.2022.11.034

22. Larsson G, Holgers KM. Fast-track care for patients with suspected hip fracture. Injury. Published online 2011. doi:10.1016/j.injury.2011.01.001

23. Larsson G, Strömberg RU, Rogmark C, Nilsdotter A. Prehospital fast track care for patients with hip fracture: Impact on time to surgery, hospital stay, post-operative complications and mortality a randomised, controlled trial. Injury. Published online 2016. doi:10.1016/j.injury.2016.01.043

24. Thomas S, Ord J, Pailthorpe C. A study of waiting time for surgery in elderly patients with hip fracture and subsequent in-patient hospital stay.

